# Modeling the Impact of Exposed Cases in a Hantavirus Outbreak on a Cruise Ship

**DOI:** 10.64898/2026.05.08.26352718

**Authors:** Jiaming Cui

**Affiliations:** Department of Computer Science, Virginia Tech

## Abstract

The emergence of a hantavirus variant aboard a commercial cruise ship presents a significant public health concern. This study develops a discrete-time stochastic Susceptible-Exposed-Infectious-Recovered-Dead model to estimate transmission dynamics, hidden exposed infections, and outbreak risk among passengers and crew. Epidemiological parameters and latent disease states were inferred using an Ensemble Adjustment Kalman Filter calibrated to reported case data from WHO and ECDC situation reports. The estimated basic reproduction number was 2.76, with a 95% confidence interval of 2.52-2.99, indicating substantial potential for sustained onboard transmission before strict quarantine measures. Simulations further suggest that several exposed individuals may remain unidentified during the early outbreak phase, creating a hidden reservoir that symptom-based surveillance alone may fail to detect. These findings highlight the importance of rapid surveillance, widespread testing, targeted quarantine, and active monitoring of exposed individuals in confined travel settings. The proposed modeling framework can support timely outbreak assessment and intervention planning for infectious-disease events in similarly dense and spatially constrained populations.

## 1 Introduction

The recent emergence and rapid spread of a hantavirus variant aboard a commercial cruise ship present a distinct and urgent public health challenge [1, 2]. Cruise ships are densely populated, spatially constrained environments in which passengers and crew share dining areas, recreational facilities, corridors, cabins, and service spaces over prolonged periods [3, 4, 5, 6]. Although confirmed symptomatic cases can be isolated rapidly once identified, transmission may continue if a substantial number of exposed individuals remain undetected [7, 8, 9]. This concern is especially important for pathogens with a long interval between exposure and symptom onset, as infected individuals may remain outside the case-identification system for an extended period before clinical signs appear, and may further spread the disease after disembarkation into larger communities [4, 10, 11].

Despite the central importance of exposed individuals, little is known about their number, timing, and contribution to outbreak growth in this setting. Confirmed case counts provide information only after infection has progressed to detectable illness or diagnostic confirmation, whereas the exposed population remains largely hidden [12, 13]. This gap limits the ability to determine whether observed cases represent isolated events or the visible portion of a larger, delayed epidemic process. In a finite and highly connected population such as a cruise ship, even a modest underestimation of the exposed pool may lead to substantial errors in projected outbreak size and in the evaluation of quarantine policies.

Therefore, this paper develops a mathematical modeling framework to estimate the hidden burden and transmission consequences of exposed infections aboard the vessel [14, 15, 16, 17]. We propose a customized SEIRD model [18] that divides the population into susceptible, exposed, infectious, recovered, and deceased compartments, with particular emphasis on the transition from exposure to symptomatic infection. This framework allows us to represent the delay between infection and detection and to evaluate how unobserved exposed individuals may sustain transmission before they are recognized as confirmed cases.

By simulating outbreak trajectories under different assumptions, we estimate that several exposed infections may remain unidentified during the early phase of the outbreak. We further estimate the basic reproduction number, R0, to be 2.76, with a 95% confidence interval of 2.52-2.99, which is similar to previous results on other Hantavirus outbreak [19]. Because R0 exceeds 1, this finding suggests the potential for sustained transmission within the cruise-ship setting. This estimate should not be directly generalized to land-based communities, where population density, contact structure, and exposure patterns may differ substantially. Nevertheless, an R0 greater than 1 highlights the need for rapid surveillance, targeted quarantine, and further assessment of exposed individuals. Overall, this study provides an initial modeling framework for hantavirus outbreak assessment in spatially constrained populations and may support timely public health decision-making in similar high-density settings.

## 2 Methods

### 2.1 Stochastic SEIRD Transmission Model

To capture the transmission dynamics of the Hantavirus variant within the closed environment of the cruise ship, we formulated a discrete-time, stochastic Susceptible-Exposed-Infectious-Recovered-Dead (SEIRD) compartmental model as shown in Figure 1. We assume that transmission occurs exclusively through contact with individuals in the infectious compartment, denoted by *I*. The exposed compartment *E* represents individuals who have been infected but have not yet developed symptoms, whereas the infectious compartment *I* represents symptomatic and documented cases.

**Figure 1:**
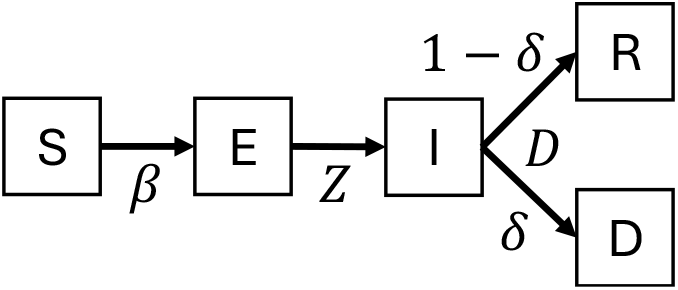
The diagram of the SEIRD model

Let *S*_*t*_, *E*_*t*_, *I*_*t*_, *R*_*t*_, and *D*_*t*_ denote the numbers of susceptible, exposed, infectious, recovered, and deceased individuals on day *t*, respectively, and let *N*_*t*_ = *S*_*t*_ + *E*_*t*_ + *I*_*t*_ + *R*_*t*_ + *D*_*t*_. The corresponding mean-field discrete-time dynamics are given by

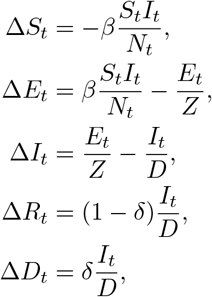

where ∆*X*_*t*_ = *X*_*t*+1_ −*X*_*t*_, *β* is the transmission rate, *Z* is the average duration of the exposed period, *D* is the average duration of the infectious period, and *δ* is the case fatality rate. To account for stochasticity in transmission and disease progression within a relatively small, confined population, the daily transition counts are modeled as Poisson random variables. Specifically,

1. **New exposures** (*U*_1,*t*_): New infections are driven by the transmission rate *β* and contact between susceptible and infectious individuals: 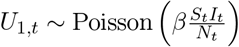.
2. **Symptom onset / documentation** (*U*_2,*t*_): Progression from the exposed compartment to the infectious compartment is governed by the average exposed period 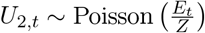.
3. **Recoveries and fatalities** (*U*_3,*t*_, *U*_4,*t*_): Removal from the infectious compartment is governed by the average infectious period *D*, with outcomes determined by the case fatality rate *δ* 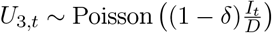, 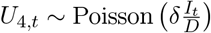.

The stochastic SEIRD model can therefore be written as the following set of daily update equations:

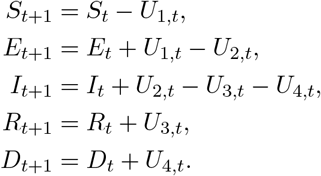

### 2.2 Parameter Inference

Epidemiological data for the cruise ship outbreak were obtained from official World Health Organization (WHO) situation reports. The dataset includes daily counts of confirmed cases, recoveries, and fatalities. Parameter values and admissible ranges were informed by previous hantavirus modeling studies, as well as publicly available WHO and European Centre for Disease Prevention and Control (ECDC) reports. The parameter ranges and estimated values are summarized in Table 1.

**Table 1:** List of parameters.

| Parameter | range | estimated value (95% CI) |
| --- | --- | --- |
| Transmission rate ($\beta$, days<sup>-1</sup>) | [0.1,0.5] [19] | 0.23 (0.22,0.25) |
| Latency period ($Z$, days) | [6,12] [14] | 9.12 (8.76,9.48) |
| Infectious period ($D$, days) | [7,14] [1, 2] | 11.52 (11.06,11.97) |
| fatality rate ($\delta$) | [0.3,0.5] [20] | 0.36 (0.35,0.38) |

For model calibration, we used the Ensemble Adjustment Kalman Filter (EAKF) to infer latent epidemiological states and estimate key model parameters. The EAKF is a recursive Bayesian data assimilation method that is well suited for nonlinear infectious disease models. By sequentially assimilating daily reported incidence data from WHO/ECDC reports into the stochastic SEIRD framework, the EAKF dynamically estimates unobserved state variables, such as the true number of exposed individuals, while simultaneously updating the posterior distributions of epidemiological parameters. Specifically, we fitted the cumulative number of transitions from the exposed compartment *E* to the infectious compartment *I* to the officially reported cumulative case counts. For the simulations, the EAKF was run for 10 iterations using an ensemble size of 300, following previous studies [21, 22].

## 3 Results

### 3.1 Model fitting

Figure 2 compares the cumulative reported cases with the simulated trajectories generated by our model over the course of the outbreak, from April 1 to May 7. As shown in the figure, the distribution of stochastic simulations closely captures the observed epidemiological data, indicated by the red crosses. In particular, the model reproduces both the slow accumulation of documented cases in early April and the subsequent stepwise increases observed in late April. The strong agreement between the reported case counts and the simulated distributions suggests that the model-inference framework can reliably reconstruct the underlying transmission dynamics and capture the observed variability within the enclosed cruise-ship environment. The estimated parameter values are listed in Table 1.

**Figure 2:**
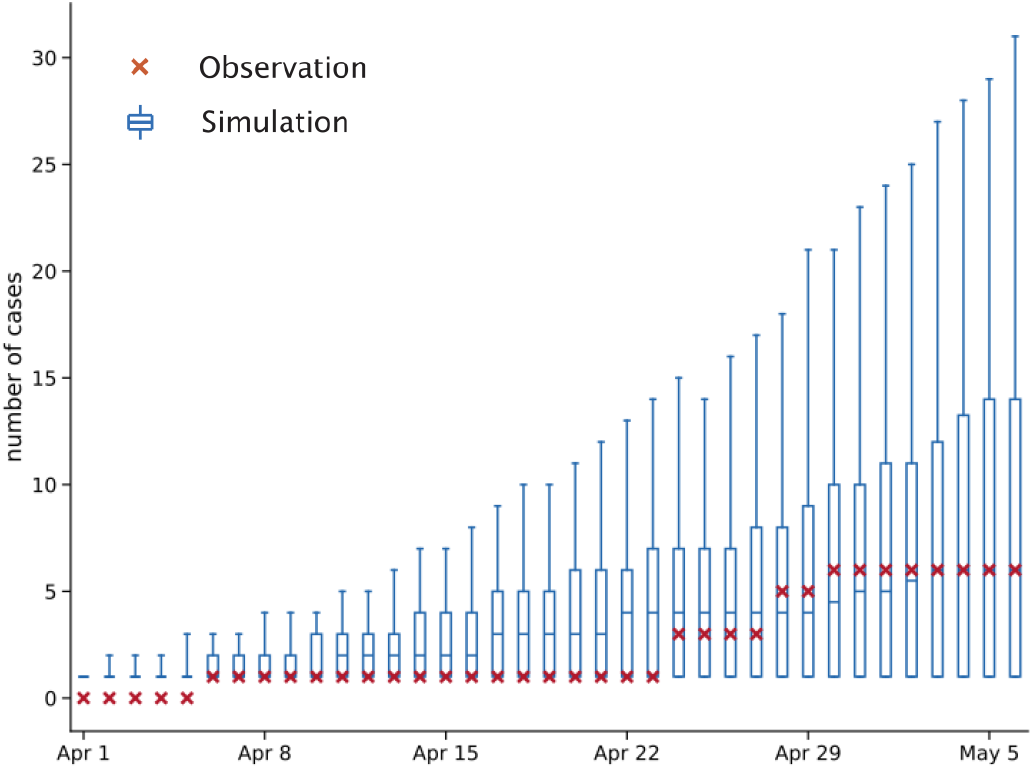
Simulated number of identified cases on board. The blue box-and-whisker plots indicate the median, interquartile range, and 95% simulation intervals.

### 3.2 Reproductive number estimation

To further evaluate the transmission potential of the Hantavirus variant in the cruise ship environment, we estimated the basic reproductive number, *R*_0_, before the implementation of strict cabin quarantine measures. As shown in Figure 3A, the estimated *R*_0_ distribution has a mean of 2.76, with a 95% confidence interval of 2.52–2.99. This elevated *R*_0_ indicates substantial potential for onboard transmission prior to quarantine.

**Figure 3:**
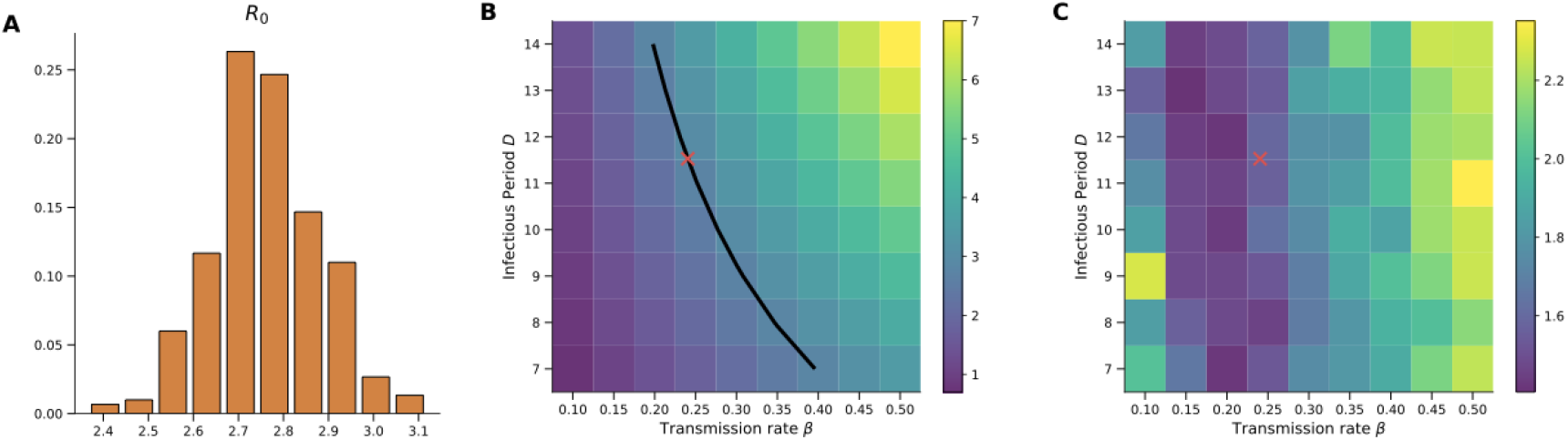
Sensitivity and identifiability analysis of estimated R0.

We further conducted sensitivity analyses to assess the identifiability of key epidemiological parameters within our model-inference framework. Specifically, we evaluated the effects of varying the transmission rate, *β*, and the infectious period, *D*, on the effective reproductive number and overall model fit, while holding all other parameters fixed at their mean estimates, as in Figure 2. In Figure 3B, the black solid line denotes *R*_0_ = 2.76, and the red cross marks the mean estimates of *β* and *D* inferred by the model.

To determine the optimal parameter combination, we calculated the root mean square error (RMSE) between model simulations and ground-truth observations across different values of the transmission rate, *β*, and the infectious period, *D*. As shown in Figure 3C, we discretized the parameter space into an 8 × 9 grid, performed simulations at each grid point, and computed the corresponding RMSE. The heat map shows that the lowest-RMSE region is strongly centered around the best-fit parameter values, *β* ≈ 0.24 and *D* ≈ 11.52, indicated by the red cross. The RMSE increases substantially as the parameter values move away from this optimum, supporting the identifiability of these parameters under the SEIRD model structure and the available daily incidence observations from the cruise ship outbreak.

### 3.3 Missing exposed cases

We further compared the number of identified cases with the number of active exposed cases. Since exposed individuals may subsequently transition to identified cases, we focus here on active exposed cases, defined as individuals who remain in the exposed state and have not yet progressed to the symptomatic or otherwise identifiable stage. As shown in Figure 4, several exposed cases remain unidentified, revealing a substantial reservoir of unisolated exposed passengers. This finding underscores that relying solely on symptom-based surveillance can severely underestimate the true scale of infection, highlighting the urgent need for active surveillance and widespread testing to identify and isolate exposed individuals before further transmission occurs.

**Figure 4:**
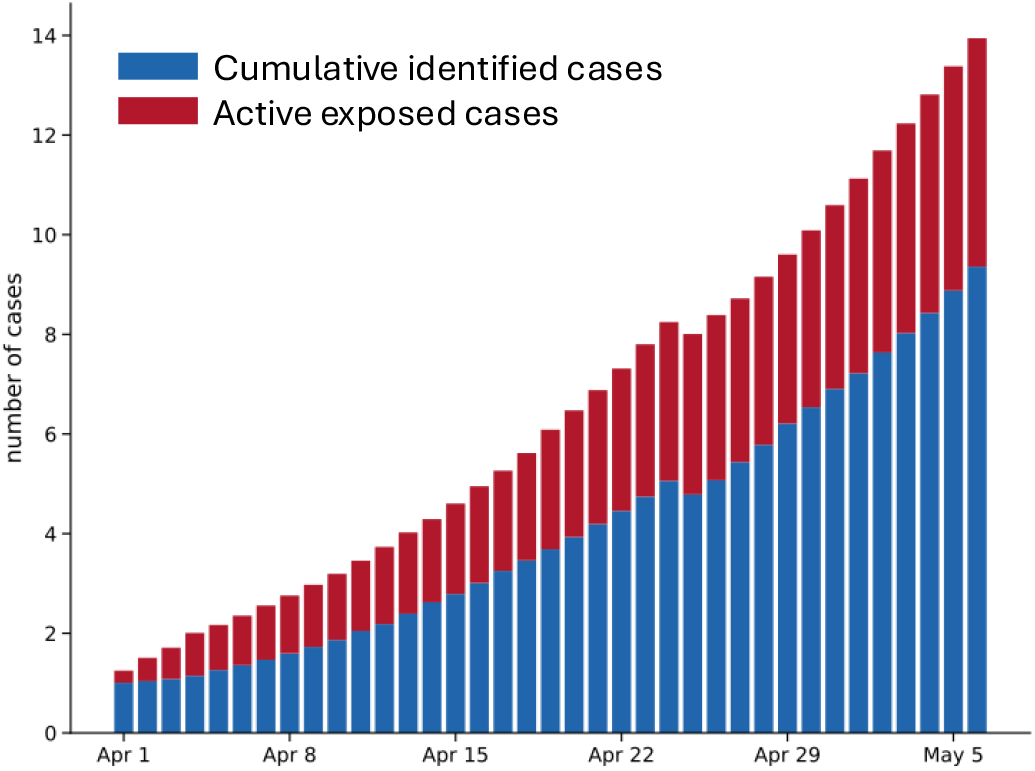
Comparison between identified cases and active exposed cases.

## 4 Discussion

This study presents an initial analysis of the transmission dynamics of a Hantavirus variant within the distinctive enclosed environment of a cruise ship. By examining a recent outbreak aboard a commercial vessel, the study addresses a timely public health concern and provides practical insights into the management of acute infectious-disease events in high-density, confined settings.

Our results indicate that the basic reproduction number, *R*_0_, in the cruise ship environment is substantially greater than 1, suggesting that sustained onboard transmission may occur in the absence of stringent control measures. The simulation results further reveal a considerable population of individuals in the exposed compartment who have not yet been detected or isolated. Because these latent or pre-symptomatic infections may lack clear clinical manifestations, such individuals can remain unidentified and may contribute to continued transmission, thereby increasing the potential scale of the outbreak. Consequently, the timely identification, monitoring, and tracing of exposed individuals remains a critical and challenging component of effective epidemic control.

Despite its value for risk assessment, this study has several limitations. The current model relies on established biological characteristics and epidemiological assumptions derived from known Hantavirus strains. However, the infectiousness, latency period, and transmission pathways of this emerging variant remain incompletely characterized. As additional epidemiological data become available, particularly data describing the variant-specific natural history and transmission patterns, the proposed model-inference framework can be further refined, calibrated, and validated.

Overall, this study underscores the substantial transmission risk posed by this Hantavirus variant in enclosed and densely populated environments. The findings provide theoretical insight into outbreak dynamics and offer practical guidance for cruise operators and public health authorities. In particular, they may support the design of future epidemic-response strategies, the optimization of early-screening and surveillance protocols, and the implementation of measures to protect the health of passengers and crew.

## Data Availability

This is a retrospective study with only public dataset

